# The Influence of NIHSS and Onset-to-Angiography Time on Neurological Outcomes Among Patients with Acute Ischemic Stroke Receiving Endovascular Thrombectomy-A Retrospective Observation Study

**DOI:** 10.1101/2025.01.29.25321366

**Authors:** Kuei-Ming Lin, Shuang-Yu Lu, Hong-Mo Shih, Tai-Yi Hsu, Fen-Wei Huang, Shih-Hao Wu, Sheng-Ta Tsai, Chien-Yu Liu, Chin-Han Lin

**Affiliations:** Department of Emergency Medicine, China Medical University Hospital, China Medical University, Taichung, Taiwan; School of Medicine, China Medical University, Taichung, Taiwan; College of Medicine, China Medical University, Taichung, Taiwan; Department of Neurology, China Medical University Hospital, China Medical University, Taichung, Taiwan; Department of Bioinformatics and Medical Engineering, Asia University, Taichung, Taiwan; Ph.D. Program for Aging, College of Medicine, China Medical University, Taichung, Taiwan

**Keywords:** Large vessel occlusion (LVO), endovascular thrombectomy (EVT)

## Abstract

**Background:** Acute ischemic stroke is a life-threatening condition requiring urgent intervention. For patients with large vessel occlusion (LVO), timely endovascular thrombectomy (EVT) is critical. This retrospective observational study investigates the impact of the National Institute of Health Stroke Scale (NIHSS) and onset-to-angiography time (OAT) on one-month neurological outcomes in acute ischemic stroke patients.

**Methods:** A total of 300 acute ischemic stroke patients with LVO who underwent EVT at a tertiary medical center in Taiwan from Jan. 2015 to Dec. 2018 were included. The NIHSS-time score was calculated by multiplying the pre-EVT NIHSS score by OAT. Favorable outcomes were defined as a modified Rankin Scale (mRS) ≤ 2 at one month. Multivariate logistic regression was used to identify the association between NIHSS-time scores and outcomes, adjusting for confounders.

**Results:** Patients with a NIHSS-time score < 4093 had significantly higher odds of achieving favorable outcomes (adjusted OR = 2.94, 95% CI = 1.56–5.56, p < 0.001). This highlights the importance of early EVT and initial stroke severity assessment in predicting recovery.

**Conclusion:** This study demonstrates that lower NIHSS-time scores are strongly associated with favorable neurological outcomes. Patients with a NIHSS-time score less than 4093 had a significantly higher ratio of favorable neurological function after one month. The NIHSS-time score offers a practical tool for prognostic evaluation, emphasizing the urgency of EVT in acute ischemic stroke patients. However, further studies are needed to validate these findings and explore their generalizability.

## Introduction

Acute ischemic stroke is a life-threatening medical condition that demands prompt intervention in the ED to minimize brain damage and improve patient outcomes. Early reperfusion therapy, such as intravenous thrombolysis and endovascular thrombectomy, has been shown to significantly enhance recovery and reduce long-term disability in stroke patients [1–6]. In Taiwan, cerebrovascular disease ranks as the fourth leading cause of death across all age groups, with ischemic stroke accounting for approximately 70% of all stroke cases [7].

The management of AIS in Taiwan involves a well-coordinated system where emergency medical technicians (EMTs) transport patients to the nearest healthcare facility capable of administering appropriate treatments. Patients who are candidates for both intravenous thrombolysis and EVT are managed under a “drip and ship” strategy [8–9], where initial thrombolysis is administered at the nearest hospital before transferring the patient to EVT-capable facility. Conversely, patients eligible solely for EVT are directly transported to a specialized hospital equipped to perform endovascular thrombectomy [10].

Timely reperfusion is critical for both intravenous thrombolysis and endovascular thrombectomy to ensure optimal neurological outcomes. However, the “drip and ship” strategy, while beneficial in certain contexts, can inadvertently increase the onset-to-angiography time (OAT) for patients with large vessel occlusion (LVO), potentially delaying crucial intervention [11].

The NIHSS is a widely used clinical tool to assess stroke severity and predict outcomes. Higher NIHSS scores correlate with more severe neurological deficits and poorer prognoses. The combination of NIHSS scores with OAT, referred to as the NIHSS-time score, provides a comprehensive measure that integrates both the severity of the stroke and the timeliness of intervention, which may be pivotal in predicting functional outcomes [12–13].

This current single-center retrospective cohort study aims to assess the influence of the NIHSS-time score on the neurological outcomes of patients with acute ischemic stroke undergoing endovascular thrombectomy. By analyzing data from a tertiary medical center in Taiwan, this study seeks to elucidate the prognostic significance of the NIHSS-time score and highlight the critical importance of minimizing OAT to enhance recovery in AIS patients.

## Methods

### Study design and setting

This retrospective observational study was conducted in the ED of China Medical University Hospital (CMUH), a prominent urban tertiary referral hospital located in Taichung City, Taiwan. The hospital is a major healthcare provider, featuring approximately 2,200 general ward beds and 130 intensive care unit (ICU) beds. In 2018, the ED alone received over 160,000 patient visits. China Medical University Hospital is equipped to provide comprehensive acute stroke care, including both intravenous thrombolysis and endovascular therapy.

Upon patient arrival at the ED, an acute stroke protocol is immediately activated during triage, ensuring rapid and systematic management of stroke cases. All patients receive standard care in accordance with the guidelines set forth by the American Heart Association, the American Stroke Association and the Taiwan Stroke Society [14–15]. Diagnostic imaging, including contrast-enhanced brain computed tomography with perfusion studies and three-dimensional vascular reconstructions, is performed concurrently. Two experienced neurologists independently review these images to confirm the diagnosis and evaluate the suitability and safety of EVT. Eligible patients received EVT within 80 minutes after arrival. To ensure accurate follow-up, the neurological disability status of patients was tracked by the case managers of the Stroke Center at China Medical University Hospital. These case managers conducted follow-up assessments, either in person or via telephone, to determine the mRS scores one month after discharge. This systematic follow-up allowed for consistent and reliable measurement of patient outcomes. The study was approved by the Institutional Review Board (IRB) of China Medical University Hospital (CMUH110-REC3-189).

### Study population

This study included all patients presenting with symptoms of acute ischemic stroke who were either directly admitted to the ED of China Medical University Hospital or referred from other hospitals between January 2015 and December 2018. China Medical University Hospital, located in Taichung City, Taiwan, serves as a major tertiary referral center, providing specialized care including intravenous thrombolysis and endovascular thrombectomy for stroke patients.

The initial analysis encompassed a comprehensive review of patient data sourced from the hospital’s electronic medical record system. The inclusion criteria were broad, capturing all individuals exhibiting stroke symptoms to ensure a representative sample of the acute ischemic stroke population. The patient selection process is shown in Figure 1. The exclusion criteria for this study were explicitly defined to maintain a clear focus on the acute ischemic stroke population receiving EVT. Patients with hemorrhagic stroke and refusal of EVT treatment were excluded if they met any of the following conditions:

**Figure 1.**
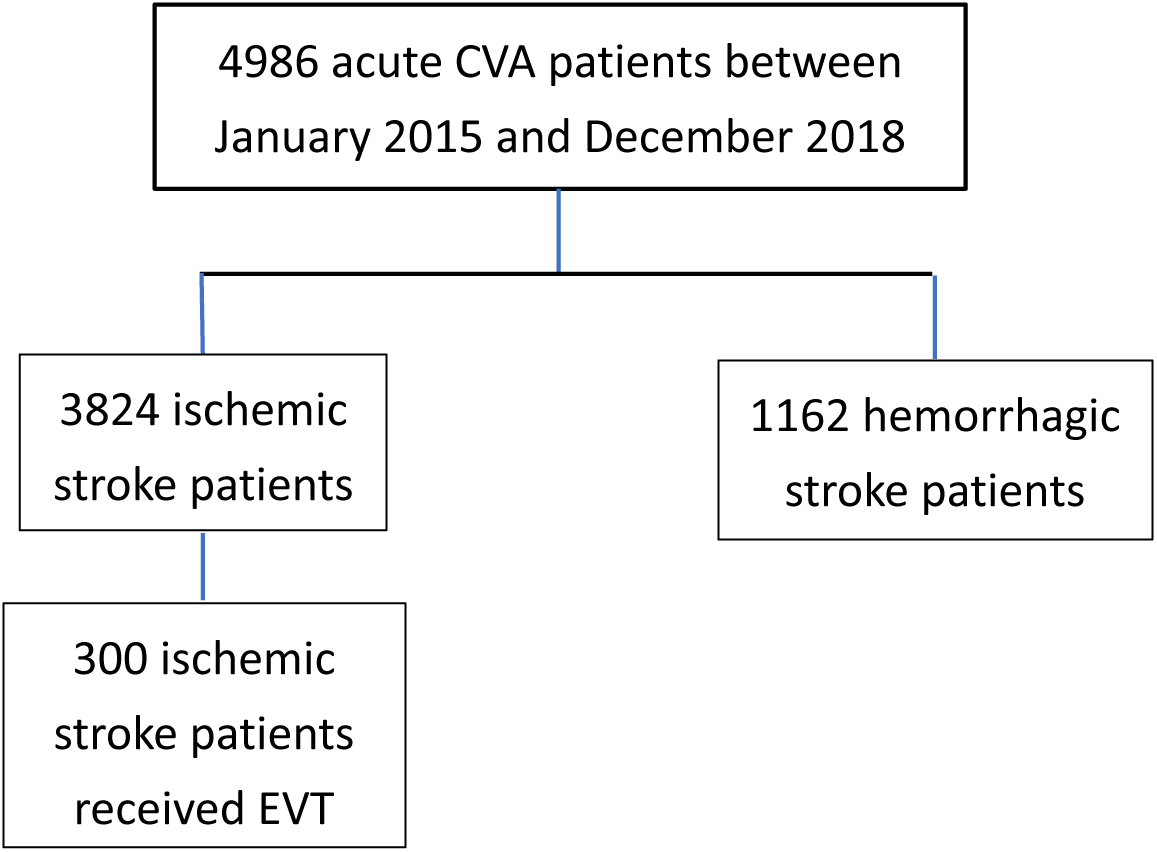
Patient enrollment. CVA = cerebrovascular accident, EVT = endovascular thrombectomy.

### Variables and data collection

Relevant variables and data collection recorded retrospectively included demographic data such as age and gender. Comprehensive medical histories were recorded, including comorbid conditions such as hypertension, diabetes mellitus, atrial fibrillation, and previous stroke events. Initial and discharge NIHSS scores were also collected, with the NIHSS being utilized at both admission and discharge to assess the severity of neurological impairment.

Onset-to-angiography time was a crucial variable in this study. OAT was defined as the interval, measured in minutes, from the time the patient was last known to be well to the time of reperfusion during EVT, and meticulously documented. The NIHSS-time score was calculated to integrate both the severity of the stroke and the timeliness of treatment. This score was computed by multiplying the NIHSS score obtained prior to EVT by the OAT. Todo et al. [13] employed the NIHSS-time score in their retrospective single-center analysis, demonstrating its utility in predicting outcomes after endovascular therapy in acute ischemic stroke patients

Finally, the Modified Rankin Scale (mRS) was used to evaluate the functional outcomes of patients one-month post-discharge. This scale, ranging from 0 (no symptoms) to 6 (death), provided a standardized measure of neurological disability and recovery.

### Outcome measurement

The primary outcome of this study was the functional outcome of patients at one-month post-discharge. To assess neurological function, we employed the modified Rankin Scale (mRS), a widely recognized and validated tool for measuring the degree of disability or dependence in daily activities of stroke survivors. The mRS scores range from 0 to 6, with each level representing a different severity of disability. For the purposes of this study, functional outcomes were dichotomized into favorable (mRS: 0-2) and unfavorable (mRS: 3-6) categories.

### Statistical analysis

To analyze the correlation between various clinical variables and neurological functional outcomes, the study population was divided into two groups based on their neurological disability status at one-month post-discharge, as measured by the modified Rankin Scale (mRS). Patients were categorized into those with favorable outcomes (mRS 0-2) and those with unfavorable outcomes (mRS 3-6).

Differences between two groups were analyzed through chi-square test for categorical variables and independent-sample *t* testing for continuous variables. Univariate and multivariate analysis were used to identify the variables correlation with neurological functional outcome.

Logistic regression was used to model the probability of achieving a favorable outcome (mRS 0-2) based on the independent variables. Odds ratios (ORs) with 95% confidence intervals (CIs) were calculated to quantify the strength of associations. Variables with p-values less than 0.05 were considered statistically significant.

To assess the predictive accuracy of the NIHSS-time score on neurological outcomes, a receiver operating characteristic (ROC) curve was constructed. The area under the curve (AUC) was calculated to quantify the overall ability of the NIHSS-time score to discriminate between patients with favorable and unfavorable outcomes. Youden’s J statistic was used to select the optimum cut off for continuous variables.

All statistical assessments were two-tailed testing. Statistical significance was defined as p<.05. All statistical analyses were processed with SAS (version 9.4; SAS Institute, Cary, NC).

## Results

### Patient population

A total of 4,986 patients presenting with acute cerebrovascular accidents (CVA) were transported or referred to China Medical University Hospital between January 2015 and December 2018. Of these, 1,162 patients were diagnosed with intracranial hemorrhage and were excluded from further analysis. The remaining 3,824 patients were diagnosed with ischemic stroke. Among these ischemic stroke patients, 300 underwent endovascular thrombectomy (EVT) (Figure 1).

### Neurological outcomes

Out of the 300 patients who received EVT, 65 patients achieved favorable neurological outcomes (mRS 0-2), while 235 patients had unfavorable outcomes (mRS 3-6). The group with favorable outcomes had a mean age of 63.52 ± 11.91 years, which was significantly younger than the unfavorable outcome group, with a mean age of 69.65 ± 11.79 years (p < 0.001). The favorable outcome group also had a lower prevalence of diabetes mellitus (35.38% vs. 50.64%, p = 0.029) and higher hyperlipidemia (32.31% vs. 14.89%, p = 0.001), as well as lower initial NIHSS scores (15.45 ± 5.49 vs. 20.35 ± 6.85, p < 0.001) and NIHSS-OAT scores (4847.0 ± 3484.1 vs. 7397.6 ± 6302.9, p < 0.001). Additionally, a higher proportion of males was observed in the favorable outcome group (72.31% vs. 54.04%, p = 0.008) (Table 1).

**Table 1.**
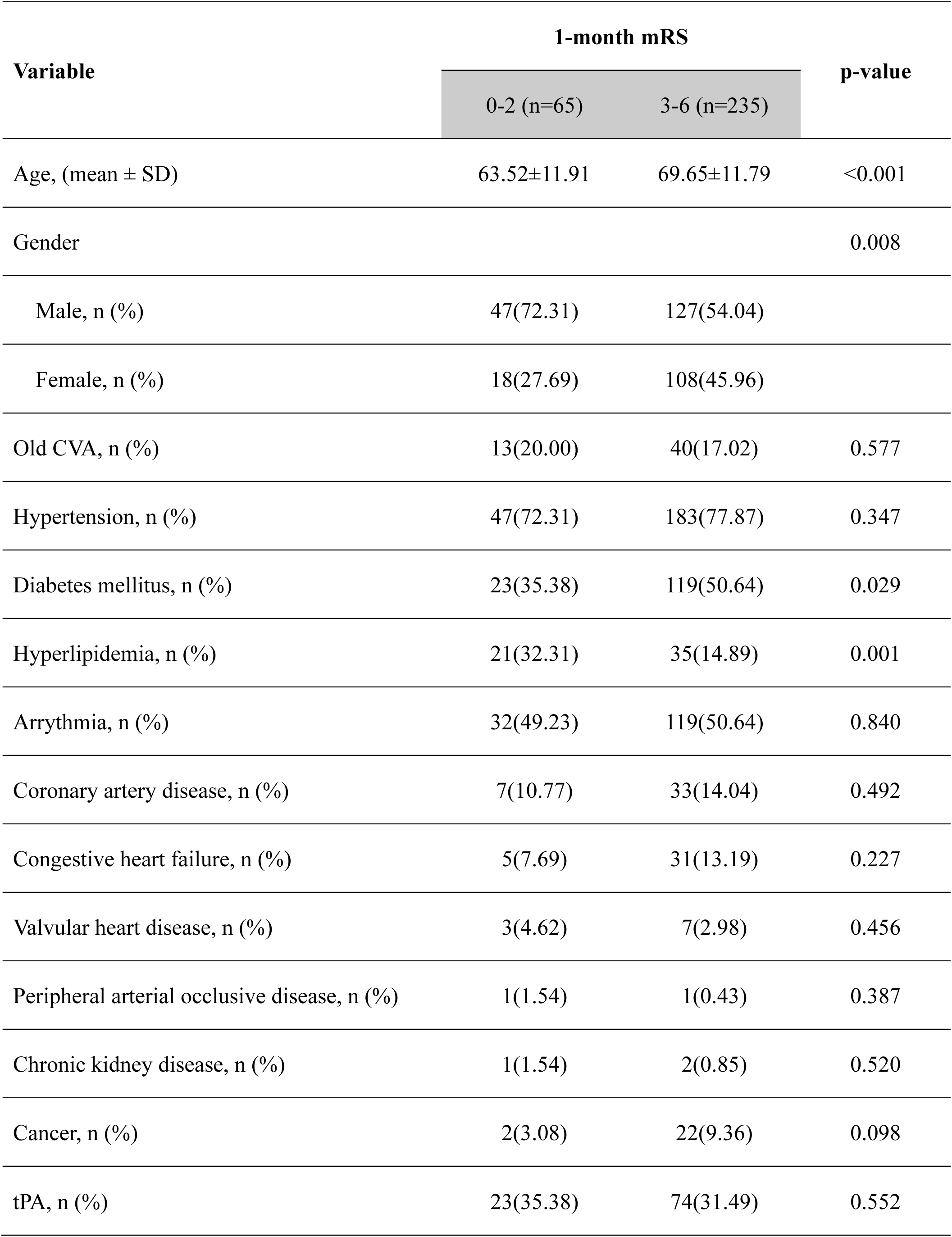
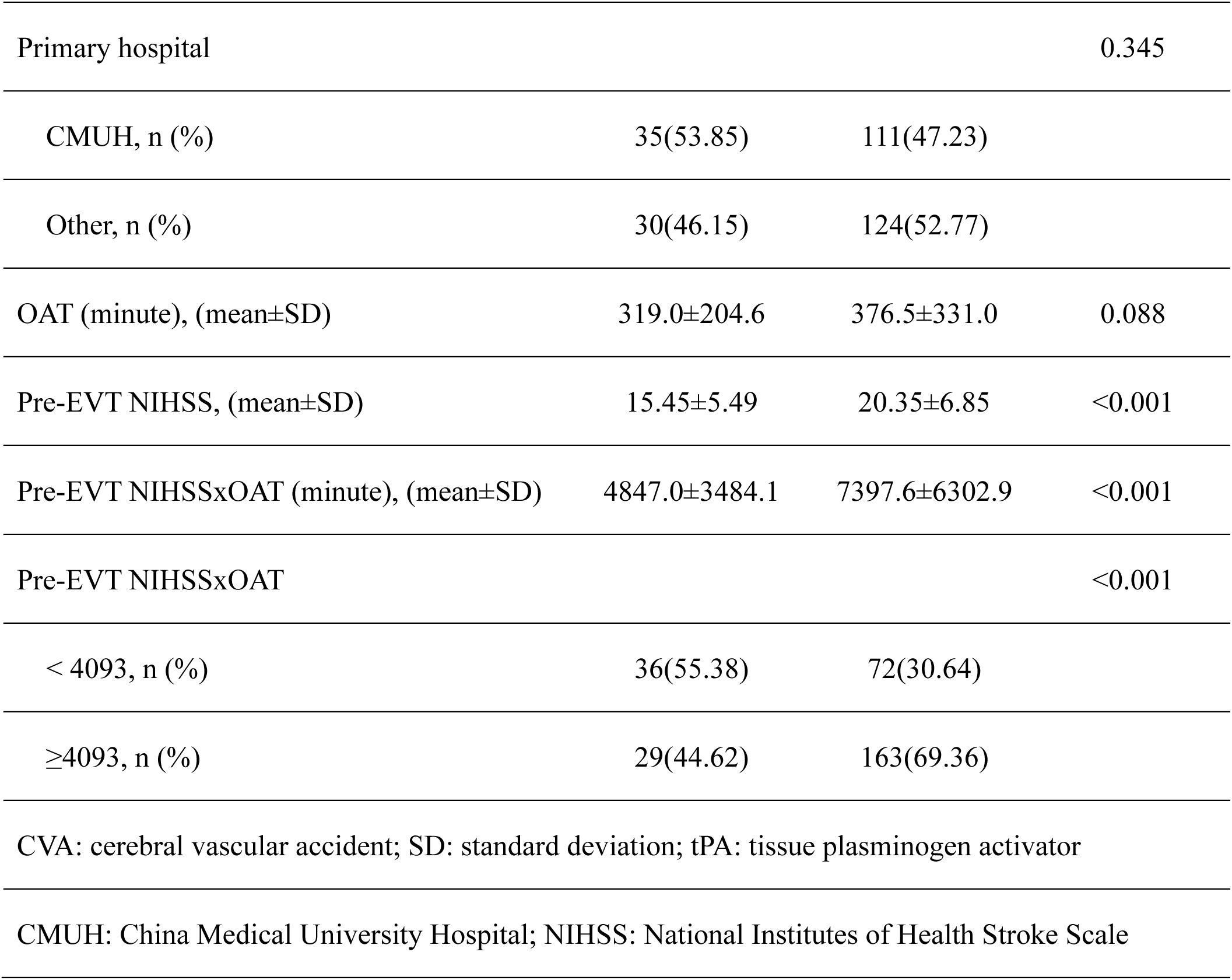
Demographic data of different neurological functional outcomes.

| Variable | 1-month mRS |  | p-value |
| --- | --- | --- | --- |
|  | 0-2 (n=65) | 3-6 (n=235) |  |
| Age, (mean ± SD) | 63.52±11.91 | 69.65±11.79 | <0.001 |
| Gender |  |  | 0.008 |
| Male, n (%) | 47(72.31) | 127(54.04) |  |
| Female, n (%) | 18(27.69) | 108(45.96) |  |
| Old CVA, n (%) | 13(20.00) | 40(17.02) | 0.577 |
| Hypertension, n (%) | 47(72.31) | 183(77.87) | 0.347 |
| Diabetes mellitus, n (%) | 23(35.38) | 119(50.64) | 0.029 |
| Hyperlipidemia, n (%) | 21(32.31) | 35(14.89) | 0.001 |
| Arrhythmia, n (%) | 32(49.23) | 119(50.64) | 0.840 |
| Coronary artery disease, n (%) | 7(10.77) | 33(14.04) | 0.492 |
| Congestive heart failure, n (%) | 5(7.69) | 31(13.19) | 0.227 |
| Valvular heart disease, n (%) | 3(4.62) | 7(2.98) | 0.456 |
| Peripheral arterial occlusive disease, n (%) | 1(1.54) | 1(0.43) | 0.387 |
| Chronic kidney disease, n (%) | 1(1.54) | 2(0.85) | 0.520 |
| Cancer, n (%) | 2(3.08) | 22(9.36) | 0.098 |
| tPA, n (%) | 23(35.38) | 74(31.49) | 0.552 |
| Primary hospital |  |  | 0.345 |
| CMUH, n (%) | 35(53.85) | 111(47.23) |  |
| Other, n (%) | 30(46.15) | 124(52.77) |  |
| OAT (minute), (mean±SD) | 319.0±204.6 | 376.5±331.0 | 0.088 |
| Pre-EVT NIHSS, (mean±SD) | 15.45±5.49 | 20.35±6.85 | <0.001 |
| Pre-EVT NIHSSxOAT (minute), (mean±SD) | 4847.0±3484.1 | 7397.6±6302.9 | <0.001 |
| Pre-EVT NIHSSxOAT |  |  | <0.001 |
| < 4093, n (%) | 36(55.38) | 72(30.64) |  |
| ≥4093, n (%) | 29(44.62) | 163(69.36) |  |
| CVA: cerebral vascular accident; SD: standard deviation; tPA: tissue plasminogen activator |  |  |  |
| CMUH: China Medical University Hospital; NIHSS: National Institutes of Health Stroke Scale |  |  |  |

### Multivariate Analysis

In the multivariate logistic regression analysis, several factors remained significantly correlated with favorable one-month neurological outcomes. These factors included younger age (OR 0.96, 95% CI 0.94-0.99, p = 0.004), absence of diabetes mellitus (OR 0.51, 95% CI 0.28-0.94, p = 0.030), presence of hyperlipidemia (OR 2.58, 95% CI 1.30-5.13, p = 0.006), lower pre-EVT NIHSS scores (OR 0.88, 95% CI 0.84-0.93, p < 0.001), and lower pre-EVT NIHSS-OAT scores (OR 0.99, 95% CI 0.98-0.99, p < 0.001). Gender and the use of intravenous thrombolysis (tPA) were not significantly associated with neurological outcomes in the multivariate analysis (Table 2).

**Table 2:** Univariate and multivariate logistic regression analysis of factors for favorable neurological functional outcomes.

| Variable | 1-month mRS | p-value | 1-month mRS | p-value |
| --- | --- | --- | --- | --- |
|  | Univariate |  | Multivariate |  |
|  | OR (95% CI) |  | OR (95% CI) |  |
| Age | 0.96 (0.94-0.98) | <0.001 | 0.96 (0.94-0.99) | 0.004 |
| Gender |  |  |  |  |
| Male | Ref. | - | Ref. | - |
| Female | 2.22 (1.22-4.05) | 0.009 | 1.85 (0.97-3.54) | 0.062 |
| Old CVA | 1.22 (0.61-2.45) | 0.577 |  |  |
| Hypertension | 0.74 (0.40-1.39) | 0.348 |  |  |
| Diabetes mellitus | 0.53 (0.30-0.94) | 0.030 | 0.51 (0.28-0.94) | 0.030 |
| Hyperlipidemia | 2.73 (1.45-5.13) | 0.001 | 2.58 (1.30-5.13) | 0.006 |
| Arrythmia | 0.95 (0.55-1.64) | 0.840 |  |  |
| Coronary artery disease | 0.74 (0.31-1.76) | 0.493 |  |  |
| Congestive heart failure | 0.55 (0.20-1.47) | 0.233 |  |  |
| Valvular heart disease | 1.58 (0.40-6.28) | 0.518 |  |  |
| PAOD | 3.66 (0.23-59.26) | 0.361 |  |  |
| Chronic kidney disease | 1.82 (0.16-20.41) | 0.626 |  |  |
| Cancer | 0.31 (0.07-1.34) | 0.117 |  |  |
| tPA | 1.19 (0.67-2.12) | 0.552 | 1.15 (0.59-2.24) | 0.683 |
| Hospital |  |  |  |  |
| CMUH | 1.30 (0.75-2.26) | 0.345 |  |  |
| Other | Ref. | - |  |  |
| OAT (minute) | 0.99 (0.99-1.00) | 0.185 |  |  |
| Pre-EVT NIHSS | 0.88 (0.84-0.93) | <0.001 |  |  |
| Pre-EVT NIHSSxOAT | 0.99 (0.98-0.99) | 0.001 |  |  |

### ROC curve analysis

The receiver operating characteristic (ROC) curve analysis of the pre-EVT NIHSS-OAT score on neurological outcomes revealed an area under the curve (AUC) of 0.6519, indicating moderate predictive accuracy (Figure 2). The optimal cutoff value for the pre-EVT NIHSS-OAT score was determined to be 4093, based on Youden’s J statistic. Patients with a pre-EVT NIHSS-OAT score less than 4093, regardless of their primary hospital source, had significantly better one-month neurological outcomes compared to those with higher scores (Table 3).

**Figure 2.**
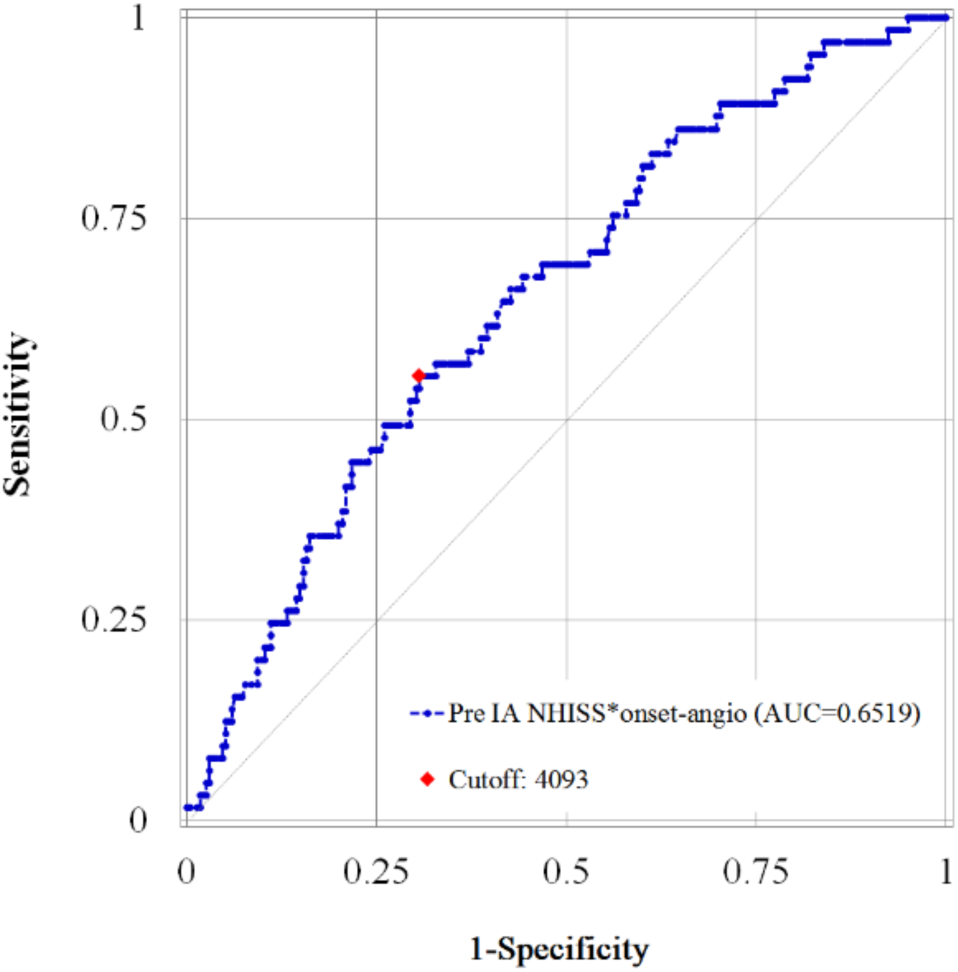
Receiver operating characteristic curve. The receiver operating characteristic (ROC) curve for pre-EVT NIHSS-time score and favorable neurological outcome. The Area under the curve is 0.6519. The best cut off pre-EVT NIHSS-time score value was 4093.

**Table 3:** Analysis of pre-EVT NIHSS-OAT score categories and hospital sources in relation to one-month neurological outcomes.

| Variable | 1-month mRS | p-value | 1-month mRS | p-value |
| --- | --- | --- | --- | --- |
|  | Univariate |  | Multivariate |  |
|  | OR (95% CI) |  | OR (95% CI) |  |
| Pre-EVT NIHSSxOAT |  |  |  |  |
| <4093 | Ref. | - |  |  |
| ≥4093 | 0.36 | <0.001 |  |  |
| Pre-EVT NIHSSxOAT<br>and hospital sources |  |  |  |  |
| <4093&CMUH | Ref. | - | Ref. | - |
| ≥4093&CMUH | 0.35 (0.16-0.78) | 0.011 | 0.28 (0.11-0.68) | 0.005 |
| <4093&other | 1.00 (0.43-2.34) | 1.000 | 0.70 (0.28-1.78) | 0.457 |
| ≥4093&other | 0.36 (0.18-0.73) | 0.004 | 0.32 (0.15-0.68) | 0.003 |

## Discussion

Our results have revealed that a lower NIHSS–time score is associated with better one-month neurological outcomes among acute ischemic stroke patients with large vessel occlusion. These findings align with those reported by Kenichi Todo et al., who similarly found that timely intervention significantly impacts stroke recovery [13]. Additionally, Karolina Skagen et al. illustrated that early recanalization serves as an independent predictor of good clinical outcomes in patients with severe ischemic stroke, particularly those with an initial NIHSS score greater than 15 [16]. This reinforces the critical importance of swift medical response. Man-Seok Park et al. reported that the “drip and ship” paradigm, which can increase the onset to groin puncture time, is associated with unfavorable functional outcomes [17]. Higher baseline NIHSS and delayed reperfusion were key predictors of hemorrhagic transformation, highlighting the need for rapid EVT in eligible patients [18]. By combining the NIHSS score with OAT, the NIHSS-time score provides a comprehensive measure that reflects both the initial neurological status and the efficiency of treatment delivery. Combining these findings, directly transporting acute stroke patients with suspected LVO to EVT-capable hospitals is crucial for improving neurological outcomes by reducing onset-to-angiography time, as highlighted by Kass-Hout et al [19]. Our results demonstrate that even patients with high NIHSS scores have the potential to achieve favorable outcomes if OAT is minimized through effective pre-hospital triage and transportation.

In our study, older age, female gender, and the presence of diabetes mellitus were correlated with poorer neurological outcomes. This is consistent with findings by Raluca Tudor et al., who identified older age and diabetes mellitus as risk factors for poor neurological outcomes, although their study found that males had worse outcomes [16]. The relationship between gender and stroke outcomes is complex. Women frequently present with atypical stroke symptoms, such as altered consciousness, fatigue, and general weakness, which can lead to misdiagnosis and delayed treatment. In Taiwan, studies have indicated that women are more likely to experience delayed hospital visits and receive less aggressive stroke interventions compared to men, contributing to poorer functional outcomes. This discrepancy highlights the need for further research to explore gender differences in stroke outcomes.

Despite the strengths of our study, several limitations should be considered. Firstly, this study’s retrospective design may cause reporting bias, particularly in the accurate documentation of OAT. Larger, multicenter cohorts are needed to confirm our results and enhance external validity. Secondly, our patients were transported by ambulances from various cities, including Taichung, Nantou, Changhua, and Yunlin, each with different dispatch policies. This variability may have influenced the OAT due to heterogeneous transportation times. Thirdly, retrospective designs make it difficult to control for confounding variables, potentially affecting causal inferences. Fourth, the study excluded patients with hemorrhagic stroke and those who refused EVT treatment. This may introduce selection bias, limiting the study’s ability to represent the full spectrum of acute ischemic stroke patients. Lastly, although the NIHSS-time score showed predictive value in a Taiwanese healthcare setting, its applicability in other regions may be limited by differences in healthcare infrastructure, EVT access, and prehospital transportation times. Future studies in diverse settings are needed to validate these findings.

## Conclusion

Patients with a NIHSS-time score less than 4093 demonstrated a favorable one-month neurological functional prognosis. Early endovascular thrombectomy in stroke patients with large vessel occlusion is critical for improving neurological outcomes. Our findings underscore the importance of efficient pre-hospital triage and direct transportation to EVT-capable hospitals to minimize treatment delays and enhance recovery prospects.

## Data Availability

The datasets generated and analyzed during the current study are not publicly available due to institutional policies and patient confidentiality agreements but are available from the corresponding author upon reasonable request. Requests must comply with ethical guidelines and require approval from the institutional review board of China Medical University Hospital.

## Acknowledgments

We would like to express our gratitude to the staffs of the Department of Emergency Medicine, and Department of Neurology and Department of Medical Imaging at China Medical University Hospital, China Medical University (Taichung, Taiwan) for their dedicated efforts in caring for the patients with ischemic stroke.

## Author contributions

Conceptualization, KML, SYL, HMS, TYH, CYL and CHL. Methodology, KML, SYL, HMS, TYH, CYL and CHL. Ethics Approval and Consent, CHL. Data collection: KML, SYL, TYH, SHW, STT, CYL and CHL. Data analysis and interpretation: HMS, TYH and FWH. Visualization, HMS, TYH, FWH. Funding acquisition, HMS and CHL. Project administration, KML, SYL, HMS, TYH, CYL and CHL. Supervision, CYL and CHL. Writing-original draft, KML and SYL. Writing-review and editing, all authors critically revised and approved the final version of the manuscript.

## Sources of Funding

The study is supported by the following grants: DMR-110-173 from China Medical University Hospital, Taichung, Taiwan.

## Disclosures

The authors declare no competing interests.

## Notes

### Competing Interest Statement

The authors have declared no competing interest.

### Clinical Trial

This study was registered retrospectively on China Medical University Hospital.

### Funding Statement

This work was supported by China Medical University Hospital, DMR-110-173. The funding source had no role in the design of the study, data collection, analysis, interpretation, or manuscript preparation. The authors or their institutions did not receive payment or services from any third party for any aspect of the submitted work.

### Author Declarations

The study was approved by the Institutional Review Board (IRB) of China Medical University Hospital (CMUH110-REC3-189).

## Reference(s)

1. Powers WJ, Derdeyn CP, Biller J, Coffey CS, Hoh BL, Jauch EC, Johnston KC, Johnston SC, Khalessi AA, Kidwell CS, et al. 2015 American Heart Association/American Stroke Association Focused Update of the 2013 Guidelines for the Early Management of Patients With Acute Ischemic Stroke Regarding Endovascular Treatment: A Guideline for Healthcare Professionals From the American Heart Association/American Stroke Association. Stroke. 2015;46:3020–3035. doi: 10.1161/STR.0000000000000074

2. Berkhemer OA, Fransen PS, Beumer D, van den Berg LA, Lingsma HF, Yoo AJ, Schonewille WJ, Vos JA, Nederkoorn PJ, Wermer MJ, et al. A randomized trial of intraarterial treatment for acute ischemic stroke. N Engl J Med. 2015;372:11–20. doi: 10.1056/NEJMoa1411587

3. Campbell BC, Mitchell PJ, Kleinig TJ, Dewey HM, Churilov L, Yassi N, Yan B, Dowling RJ, Parsons MW, Oxley TJ, et al. Endovascular therapy for ischemic stroke with perfusion-imaging selection. N Engl J Med. 2015;372:1009–1018. doi: 10.1056/NEJMoa1414792

4. Goyal M, Demchuk AM, Menon BK, Eesa M, Rempel JL, Thornton J, Roy D, Jovin TG, Willinsky RA, Sapkota BL, et al. Randomized assessment of rapid endovascular treatment of ischemic stroke. N Engl J Med. 2015;372:1019–1030. doi: 10.1056/NEJMoa1414905

5. Saver JL, Goyal M, Bonafe A, Diener HC, Levy EI, Pereira VM, Albers GW, Cognard C, Cohen DJ, Hacke W, et al. Stent-retriever thrombectomy after intravenous t-PA vs. t-PA alone in stroke. N Engl J Med. 2015;372:2285–2295. doi: 10.1056/NEJMoa1415061

6. Jovin TG, Chamorro A, Cobo E, de Miquel MA, Molina CA, Rovira A, San Roman L, Serena J, Abilleira S, Ribo M, et al. Thrombectomy within 8 hours after symptom onset in ischemic stroke. N Engl J Med. 2015;372:2296–2306. doi: 10.1056/NEJMoa1503780

7. Ministry of Health and Welfare, Taiwan. (2020). Statistics of Causes of Death. https://www.mohw.gov.tw/cp-5256-63399-2.html

8. Deguchi I, Mizuno S, Kohyama S, Tanahashi N, Takao M. Drip-and-Ship Thrombolytic Therapy for Acute Ischemic Stroke. J Stroke Cerebrovasc Dis. 2018;27:61–67. doi: 10.1016/j.jstrokecerebrovasdis.2017.07.033

9. Broderick JP, Palesch YY, Demchuk AM, Yeatts SD, Khatri P, Hill MD, Jauch EC, Jovin TG, Yan B, Silver FL, et al. Endovascular therapy after intravenous t-PA versus t-PA alone for stroke. N Engl J Med. 2013;368:893–903. doi: 10.1056/NEJMoa1214300

10. Saver JL, Fonarow GC, Smith EE, Reeves MJ, Grau-Sepulveda MV, Pan W, Olson DM, Hernandez AF, Peterson ED, Schwamm LH. Time to treatment with intravenous tissue plasminogen activator and outcome from acute ischemic stroke. JAMA. 2013;309:2480–2488. doi: 10.1001/jama.2013.6959

11. Meretoja A, Strbian D, Mustanoja S, Tatlisumak T, Lindsberg PJ, Kaste M. Reducing in-hospital delay to 20 minutes in stroke thrombolysis. Neurology. 2012;79:306–313. doi: 10.1212/WNL.0b013e31825d6011

12. Jr HPA, P H Davis ECL, Chang KC, Bendixen BH, Clarke WR, Woolson RF, Hansen MD. Baseline NIH Stroke Scale score strongly predicts outcome after stroke A report of the Trial of Org 10172 in Acute Stroke Treatment (TOAST). Neurology. 1999;53:126–131. doi: 10.1212/wnl.53.1.126

13. Todo K, Sakai N, Kono T, Hoshi T, Imamura H, Adachi H, Kohara N. National Institutes of Health Stroke Scale-Time Score Predicts Outcome after Endovascular Therapy in Acute Ischemic Stroke: A Retrospective Single-Center Study. J Stroke Cerebrovasc Dis. 2016;25:1187–1191. doi: 10.1016/j.jstrokecerebrovasdis.2016.01.027

14. Powers WJ, Derdeyn CP, Biller J, Coffey CS, Hoh BL, Jauch EC, Johnston KC, Johnston SC, Khalessi AA, Kidwell CS, et al. 2015 American Heart Association/American Stroke Association Focused Update of the 2013 Guidelines for the Early Management of Patients With Acute Ischemic Stroke Regarding Endovascular Treatment: A Guideline for Healthcare Professionals From the American Heart Association/American Stroke Association. Stroke. 2015;46:3020–3035. doi: 10.1161/STR.0000000000000074

15. Taiwan Stroke Society. (2013, September). Guidelines of Ischemic Stroke. https://www.stroke.org.tw/GoWeb2/include/getfile.php?file=f01&KeyID=20369671756644140fbfb02

16. Skagen K, Skjelland M, Russell D, Jacobsen EA. Large-Vessel Occlusion Stroke: Effect of Recanalization on Outcome Depends on the National Institutes of Health Stroke Scale Score. J Stroke Cerebrovasc Dis. 2015;24:1532–1539. doi: 10.1016/j.jstrokecerebrovasdis.2015.03.020

17. Park MS, Lee JS, Park TH, Cho YJ, Hong KS, Park JM, Kang K, Lee KB, Kim JG, Lee SJ, et al. Characteristics of the Drip-and-Ship Paradigm for Patients with Acute Ischemic Stroke in South Korea. J Stroke Cerebrovasc Dis. 2016;25:2678–2687. doi: 10.1016/j.jstrokecerebrovasdis.2016.07.015

18. Iancu A, Buleu F, Chita DS, Tutelca A, Tudor R, Brad S. Early Hemorrhagic Transformation after Reperfusion Therapy in Patients with Acute Ischemic Stroke: Analysis of Risk Factors and Predictors. Brain Sci. 2023;13. doi: 10.3390/brainsci13050840

19. Kass-Hout T, Lee J, Tataris K, Richards CT, Markul E, Weber J, Mendelson S, O’Neill K, Sednew RM, Prabhakaran S. Prehospital Comprehensive Stroke Center vs Primary Stroke Center Triage in Patients With Suspected Large Vessel Occlusion Stroke. JAMA Neurol. 2021;78:1220–1227. doi: 10.1001/jamaneurol.2021.2485

